# Exposure to perfluoroalkyl substances in a cohort of women firefighters and office workers in San Francisco

**DOI:** 10.1101/19005652

**Authors:** Jessica Trowbridge, Roy Gerona, Thomas Lin, Ruthann A. Rudel, Vincent Bessonneau, Heather Buren, Rachel Morello-Frosch

## Abstract

**Background:** Studies in male firefighters have demonstrated increased exposures to carcinogenic compounds and increased rates of certain cancers compared to the general population. Many chemicals related to these occupational exposures have been associated with breast tumor development in animal and human studies, yet, there have been no studies on women firefighters due to their low numbers in most fire departments. To address this data gap, the Women Firefighters Biomonitoring Collaborative (WFBC) created a biological sample archive and analyzed levels of perfluoroalkyl substances (PFAS) among women firefighters and office workers in San Francisco.

**Methods:** Active duty women firefighters (n=86) and office workers (n=84) were recruited from the San Francisco Fire Department and the City and County of San Francisco, respectively. Serum samples were collected and analyzed using liquid chromatography tandem mass spectrometry (LC MS/MS) to measure and compare PFAS levels between firefighters and office workers. For PFAS congeners detected in at least 70% of our study population, we examined differences in serum PFAS levels controlling for dietary, demographic and other confounders. Among firefighters, we assessed associations between occupational activities and PFAS levels.

**Results:** Eight of 12 PFAS congeners were detected at levels above the limit of detection and seven were detected in at least 70% of the study population. Four PFAS were detected in all study participants (PFNA, PFOA, PFOS, PFHxS). In regression models comparing PFAS levels by occupation and adjusting for potential confounders, firefighters had higher geometric mean (GM) concentrations of PFAS compared to office workers: 2.39 (95%CI = 1.64,3.48), 2.32 (95% CI = 1.17,4.62) and 1.26 (95% CI = 0.99, 1.59) times higher for PFHxS, PFUnDA and PFNA, respectively. In analyses limited to firefighters, PFAS levels varied by assigned position in the fire department—firefighters and officers had higher PFNA, PFOA, PFDA, and PFUnDA compared to drivers. Additionally, firefighters who reported having used firefighting foam had higher concentrations of PFOA compared firefighters who reported never having used foam.

**Conclusion:** Our study found ubiquitous exposures to PFAS among WFBC participants, with women firefighters exposed to higher levels of some PFAS compared to office workers, suggesting that some of these exposures may be occupationally related.

## Introduction

Firefighters have higher rates of some cancers compared to the general population. A meta-analysis of 32 studies found elevated rates of lymphoma, testicular, and prostate cancer among male firefighters.^1^ Additionally, a review by the International Agency for Research on Cancer (IARC) found increased cancer rates among firefighters, and designated the occupation of firefighting as “possibly carcinogenic” or “class 2b”.^2^ More recently, a study of over 19,000 United States firefighters conducted by the National Institute for Occupational Health and Safety (NIOSH) found that total time spent at fires was associated with increased lung cancer incidence and mortality, and total number of fire responses was associated with leukemia mortality.^3^ Furthermore, studies in other countries have found increased rates of several cancers among male firefighters and other first responders including brain, thyroid, bladder, kidney, prostate, testicular, breast, digestive cancers, multiple myeloma, and non-Hodgkin’s lymphoma.^4–10^ Despite mounting concern about cancer among male firefighters, few studies have assessed chemical exposures or cancer risk among women firefighters, many of whom are concerned about the potential increased risk of breast cancer and other reproductive cancers. Only two studies have examined the incidence of cancer among women firefighters. Daniels et al. found that women firefighters had higher incidence and mortality rates of breast cancer compared to the general US population, though neither effect estimate was statistically significant.^3^ A study of Florida firefighters found that women firefighters had an increase in overall cancer risk and, in particular, an increased incidence of Hodgkin’s disease and cervical and thyroid cancers compared to the general Florida population.^9^

Occupational exposures may be an important contributor to the increased risk of cancer among firefighters. Firefighters’ exposure to environmental chemicals may arise from fire suppression activities and during the salvage and overhaul phase of a fire event.^11–15^ Additionally, hazardous chemicals have been identified in fire station dust, firefighting foams, diesel emissions, contaminated fire equipment and firefighter protective gear.^16–20^ Previous studies have shown that firefighters are occupationally exposed to polycyclic aromatic hydrocarbons,^13,21–24^ formaldehyde, dioxins, polybrominated diphenyl eithers (PBDEs), organophosphate flame retardants,^25,26^ and perfluoroalkyl substances (PFAS),^19,25,27,28^ among others.^25,29,30^ Many of these chemicals have been associated with adverse health outcomes including breast cancer and breast tumor development in both animal and human studies.^31,32^

PFAS may be of particular relevance to firefighting because these compounds are used in turnout gear and are a major ingredient of some firefighting foams, such as aqueous film-forming foams (AFFF).^19,28,33^ More generally, PFAS chemicals are frequently applied to food contact paper, fabrics and furniture to make them stain, water and grease resistant.^34^ PFAS have long half-lives and bioaccumulate in the environment and human body.^35^ Because of their widespread use and persistence, they have been detected in nearly everyone tested in large biomonitoring studies.^36,37^ Biomonitoring studies within the National Health and Nutrition Examination Survey (NHANES), a nationally representative sample of the U.S. population, have found that more than 98% of people tested had multiple congeners of PFAS detected in their bodies.^36^

PFAS exposure has been linked to multiple adverse health outcomes including cancer, immune suppression, thyroid and sex hormone disruption, and decreased semen quality.^38–41^ Studies also indicate that exposure is associated with metabolic effects, ulcerative colitis and adverse effects on liver and kidney function.^42–45^

Several firefighter biomonitoring studies measured PFAS levels and found higher levels among firefighters compared to the general population.^25,27,28^ These studies, however, included few or no women. Overall, women remain underrepresented in studies of firefighters, and very little is known about the extent of their chemical exposures or occupational health risks for diseases such breast cancer.

Although women make up 5.1% of firefighters across the United States,^46^ their numbers can be higher in urban jurisdictions, including in San Francisco, which has one of the highest proportions of women firefighters (15%) in any large urban fire department in the U.S.^47^ As fire departments and other first responder professions diversify and recruit more women to their ranks, it is important to characterize chemical exposures and implications for health outcomes of particular relevance to women, such as breast cancer. To address this data gap, a partnership of firefighters, environmental health scientists, and environmental health advocates created the Women Firefighters Biomonitoring Collaborative (WFBC). The WFBC is a community-based, participatory biomonitoring project that aims to better understand how women firefighters are exposed to potential breast carcinogens, while also developing a biospecimen archive of women firefighters and office workers in San Francisco. As part of the WFBC, we conducted a cross-sectional chemical biomonitoring study to compare levels of PFAS in human serum collected from women firefighters from the San Francisco Fire Department (SFFD) and non-firefighter women who are office workers for the City and County of San Francisco. To our knowledge, this is the first biomonitoring study to measure environmental chemical exposures in an exclusively female cohort of firefighters and office workers.

## Methods

### Recruitment

Participant recruitment and sample collection took place between June 2014 and March 2015. Firefighter participants were recruited from the SFFD, via word-of-mouth, emails through the Department listserv and an article about the study in the “The Mainline,” a Department newsletter written and edited by both active and retired firefighters. Similarly, office workers, who were non-first responder employees of the City and County of San Francisco, were recruited via word-of-mouth, public meetings, health fairs, and listserv emails targeting employees of the City and County of San Francisco.

Study inclusion criteria for both firefighters and office workers included being female, over 18 years old, a full-time employee, and a non-smoker. In addition, firefighters needed to have at least five years of service with the SFFD and currently be on “active duty” (i.e. assigned to a fire station) at the time of recruitment. All participants were consented into the study following protocols approved by the Institutional Review Board of the University of California, Berkeley (# 2013-07-5512).

### Exposure assessment interview

After consent and enrollment into the WFBC study, we conducted an hour-long in-person exposure assessment interview with each participant. The interview captured demographic information, basic health information, and possible sources of PFAS exposure from occupational activities, consumer product use, and dietary factors that prior literature indicates are potential sources of PFAS exposure.^27,48,49^ Food frequency responses were converted to times per week and categorized into quartiles, tertiles, or ever/never.

### Sample collection and processing

Blood samples were collected by a certified phlebotomist in 40 mL additive-free glass tubes and transported in a cooler with ice for processing within 3 hours of collection. Serum was separated by allowing clotting at room temperature, then centrifuging at 3000 rpm for 10 minutes. Serum was aliquoted into 1.2 mL cryo-vial tubes and stored at −80 °C until analysis per standard protocols.^50^ All samples were processed and analyzed at the University of California, San Francisco.

### Laboratory analysis

Twelve PFAS (perfluorobutanoic acid, PFBA; perfluorobutane sulfonic acid, PFBuS; perfluorohexanoic acid, PFHxA; perfluorohexane sulfonic acid, PFHxS; perfluoroheptanoic acid, PFHpA; perfluorooctanoic acid, PFOA; perfluorooctane sulfonic acid, PFOS; perfluorooctane sulfonamide, PFOSA; perfluorononanoic acid, PFNA; perfluorodecanoic acid, PFDA; perfluoroundecanoic acid, PFUnDA; perfluorododecanoic acid, PFDoA) were analyzed in each serum sample (0.5 mL) using liquid chromatography-tandem mass spectrometry (LC-MS/MS). An Agilent LC1260 (Sta. Clara, CA)-AB Sciex API 5500 (Foster City, CA) platform was used in the analysis. Prior to injection into the LC-MS/MS, each sample was prepared for analysis by solid phase extraction using a Waters Oasis HLB cartridge (10 mg, 1cc). Extracted aliquots of each sample (25uL) were run in duplicates. The twelve analytes were separated by elution gradient chromatography using Phenomenex Kinetex C18 column (100 × 4.6 mm, 2.6µ) at 40°C. An electrospray ionization source operated in the negative mode was used to ionize each analyte in the mass spectrometer.

Analytes were detected in each sample by multiple reaction monitoring using two transitions per analyte. To determine the presence of each analyte retention matching (within 0.15 min) along with the peak area ratio between its qualifier and quantifier ions (within 20%) were used. Quantification of each detected analyte was done by isotope dilution method using a 10-point calibration curve (0.02-50 ng/mL) and employing two C13-labelled PFAS isotopologues. The limits of quantification for the twelve analytes range from 0.05 to 0.1 ng/mL. Analyte identification from total ion chromatograms was evaluated using AB Sciex Analyst v2.1 software while quantification of each analyte was processed using AB Sciex MultiQuant v2.02 software. Analysts were blinded to firefighter and office worker status of the serum samples during the analysis. Results were reported in ng/mL for all 170 study participants.

### Statistical analysis

We examined the distribution of each PFAS congener across the study population and then separately for firefighters and office workers and calculated summary statistics including geometric mean (GM), 95% confidence intervals (CI), and percentiles. As is common with environmental data, PFAS concentrations were non-normally distributed, thus we used non-parametric methods (Wilcoxon rank-sum test) to test unadjusted differences in PFAS concentrations between firefighters and office workers. We used lognormal regression analyses to assess differences in PFAS concentrations between firefighters and office workers, controlling for potential confounders. We then limited the analysis to firefighters and used lognormal regression analyses to explore the association between firefighter occupational activities and PFAS concentrations controlling for potential confounders. Congeners with at least a 70% detection frequency were included in the data analysis and we used maximum likelihood estimation (MLE) with the NADA R package to account for left censored data (data below the limit of detection (LOD)) for all regression models.^51^

Potential confounders were identified *a priori* based on previous literature suggesting an independent relationship with PFAS levels. We assessed the relationship between each identified variable and PFAS levels in our data and tested differences between firefighters and office workers for each potential confounder associated. Variables were included in final regression models if there was a statistically significant association with PFAS levels in our data and if the variable demonstrated a statistically significant difference between firefighters and office workers (p-value ≤ 0.05) for at least one PFAS congener.

We ran linear regression models to test the association between occupation and log-transformed PFAS concentrations controlling for age, race and ethnicity, and education (Model 1). A second model controlled for variables in Model 1 as well as consumption frequency of fish and shellfish, red meat, poultry, fast-food or take-out food, and frozen food heated in paper or cardboard packaging (Model 2). Exponentiated beta coefficients estimate the proportional change in the PFAS geometric means associated with being a firefighter compared to being an office worker and controlling for potential confounders.

We then limited our analysis to firefighters to evaluate the association between firefighter practices in the workplace with PFAS levels adjusting for age, race and ethnicity, and number of years of service with SFFD. Firefighter practices assessed included use of self-contained breathing apparatus (SCBA) during salvage and overhaul, use of firefighting foam, and the participants’ assigned position (firefighter, officer, driver). Likewise, because airport fire stations are required to stock, test and use PFAS-containing firefighting foam due to federal regulations, we included an indicator variable for those firefighters assigned to San Francisco Airport fire stations.^52^ We also examined the relationship between PFAS concentrations and the frequency of handwashing during a work shift and showering after a fire event, as well as the relationship with responding to a fire within 24 hours and one year prior to providing a biospecimen sample. Again, we exponentiated the beta coefficient to obtain the proportional change in the PFAS geometric means associated with a unit change of each dependent variable controlling for potential confounders.

Lastly, we evaluated how PFAS levels in our study population compared with those measured in the general U.S. population and in firefighters in Southern California. Specifically, we plotted the geometric mean (GM) and 95% confidence interval from the WFBC firefighter and office worker groups and compared them to adult women from the 2013-2014 cycle of the National Health and Nutrition Examination Survey (NHANES) and the Firefighter Occupational Exposure (FOX) study, in which samples were collected from a cohort of mostly male firefighters in Southern California between 2010 and 2011.^27,53,54^ WFBC firefighter and office worker levels below the LOD were replaced by the LOD reported for each congener divided by √2 to facilitate comparison of our results to the FOX and NHANES cohorts, which also used this approach (LOD/√2).

All analyses were performed in R version 3.5.2 and R Studio 1.2.1335.^55,56^

## Results

176 participants were recruited into the study. Six individuals, (three firefighters and three office workers) met the study inclusion criteria and were interviewed but did not provide a blood sample and therefore were excluded from the analysis. Our final study sample consisted of 86 firefighters and 84 office workers (N=170) (**Table 1**).

**Table 1.** Demographic characteristics of firefighters and office workers in the Women Firefighters Biomonitoring Collaborative (2014-2015)

| **Variables: mean ± SD or N(%)** | **Firefighters**<br>**N=86** | **Office workers**<br>**N=84** | **P-value<sup>a</sup>** |
| --- | --- | --- | --- |
| Age (years) | 47.5 (±4.6) | 48.3 (±10.5) | 0.38 |
| Years working for SFFD or SF City & County | 17.4 (±4.2) | 14.2 (±10.1) | 0.0005 |
| U.S. Born | 77 (89.5%) | 62 (73.8%) | 0.01 |
| **Race/ethnicity** |  |  |  |
| NH White | 40 (46.5%) | 37 (44.0%) | 0.29 |
| Latina/Hispanic | 19 (22.1%) | 13 (15.5%) |  |
| NH Asian | 11 (12.8%) | 19 (22.6%) |  |
| NH Black | 9 (10.5%) | 5 (6.0%) |  |
| NH Other | 7 (8.1%) | 10 (11.9%) |  |
| **Last grade completed** |  |  |  |
| High school or equivalent | 6 (7.0%) | 4 (4.8%) | < 0.0001 |
| Some college | 42 (48.8%) | 11 (13.1%) |  |
| Bachelors | 32 (37.2%) | 26 (31.0%) |  |
| Post Graduate | 6 (7.0%) | 43 (51.2%) |  |
| **Marital status** |  |  |  |
| Married | 37 (43.0%) | 47 (56.0%) | 0.039 |
| Widowed | 1 (1.2%) | 2 (2.4%) |  |
| Separated/Divorced | 26 (30.2%) | 11 (13.1%) |  |
| Never married | 22 (25.6%) | 24 (28.6%) |  |
| **Reported yes, ever been pregnant** | 60 (70.6%) | 52 (61.9%) | 0.26 |
| **Body mass index<sup>b</sup>** |  |  |  |
| Healthy weight | 33 (40.7%) | 43 (52.4%) | 0.13 |
| Overweight | 35 (43.2%) | 23 (28.0%) |  |
| Obese | 13 (16.0%) | 16 (19.5%) |  |

NH = Non-Hispanic
<sup>a</sup>To account for non-normally distributed variables we used Wilcoxon Rank Sum test to compare continuous variables by firefighter status and the Fisher Test for categorical variables.
<sup>b</sup> CDC guidelines for BMI classification (lb/in<sup>2</sup>): Healthy weight 18.5-24.9; Overweight 25.0-29.9, Obese >30

Firefighters and office workers had similar demographic characteristics in terms of age, and racial/ethnic make-up, while a significantly higher proportion of office workers were foreign born, married, and had higher levels of educational attainment. Firefighters had an average of seven years of service with the SFFD while office workers had an average of fourteen years of service with the City and County of San Francisco.

Occupational activities and characteristics of the firefighter group are shown in **Table 2**. The majority of participants were assigned to the position of firefighter compared to officer or driver positions, and thirteen firefighters were assigned to one of the San Francisco airport fire stations. Most firefighters reported using firefighting foam during their career as a firefighter and over half of participants reported rarely using their SCBA during salvage and overhaul activities after a fire. When asked about recent fires, most firefighters had not responded to a fire or participated in live-fire training in the 24 hours prior to their biospecimen sample collection, while the SFFD fire history data indicated that most firefighters had ten or more fires in the year prior to biospecimen collection.

**Table 2.** Occupational characteristics for WFBC firefighters 2014-15 (N = 86)

| Variables | Mean(±SD) or N(%) |
| --- | --- |
| Frequency of handwashing while at work (times/shift) | 18.8 (±12.5) |
| Frequency of handwashing while not at work (times/day) | 10.3 (±5.3) |
| Reported ever using firefighting foam | 77 (89.5%) |
| Assigned to airport fire station | 14 (16.3%) |
| Response to fire in last 24 hours<sup>a</sup> | 15 (17.4%) |
| **Number of fires responded to in year prior to sample collection (N = 66)<sup>b</sup>** |  |
| ≤9 | 17 (25.8%) |
| 10-15 | 17 (25.8%) |
| 16-19 | 15 (22.7%) |
| ≥20 | 17 (25.8%) |
| **Assignment in the SFFD** |  |
| Driver | 21 (24.4%) |
| Firefighter | 25 (29.1%) |
| Officer | 40 (46.5%) |
| **Use self-contained breathing apparatus (SCBA) during salvage/overhaul<sup>c</sup>** |  |
| Always/often | 12 (15.0%) |
| Sometimes | 14 (17.5%) |
| Rarely/never | 54 (67.5%) |

<sup>a</sup>Includes fire response and training involving live fires; <sup>b</sup>Includes firefighters who consented to give access to SFFD fire history records (n = 66); <sup>c</sup>Six firefighters reported 'do not participate' in salvage/overhaul.

### Perfluoroalkyl substance exposures

**Table 3** shows the LOD, detection frequency, GM (95%CI), and percentiles of each PFAS compound analyzed. Of the twelve PFAS we measured in participants’ serum, four congeners were not detected in any of the study participants (PFBA, PFHxA, PFHpA, PFOSA). Eight PFAS had measurable levels, seven of which were detected in at least 70% of study participants and four were detected in 100% of study participants (PFHxS, PFNA, PFOA, and PFOS). We excluded PFDoA from further analyses since it was detected in fewer than 25% of study participants. Distributional comparisons of PFAS levels between WFBC firefighters and office workers for those compounds with detection frequencies of at least 70% are shown in **Figure S1**. Levels of PFNA, PFHxS and PFUnDA were statistically significantly higher among the firefighter group compared to office workers.

**Table 3.**
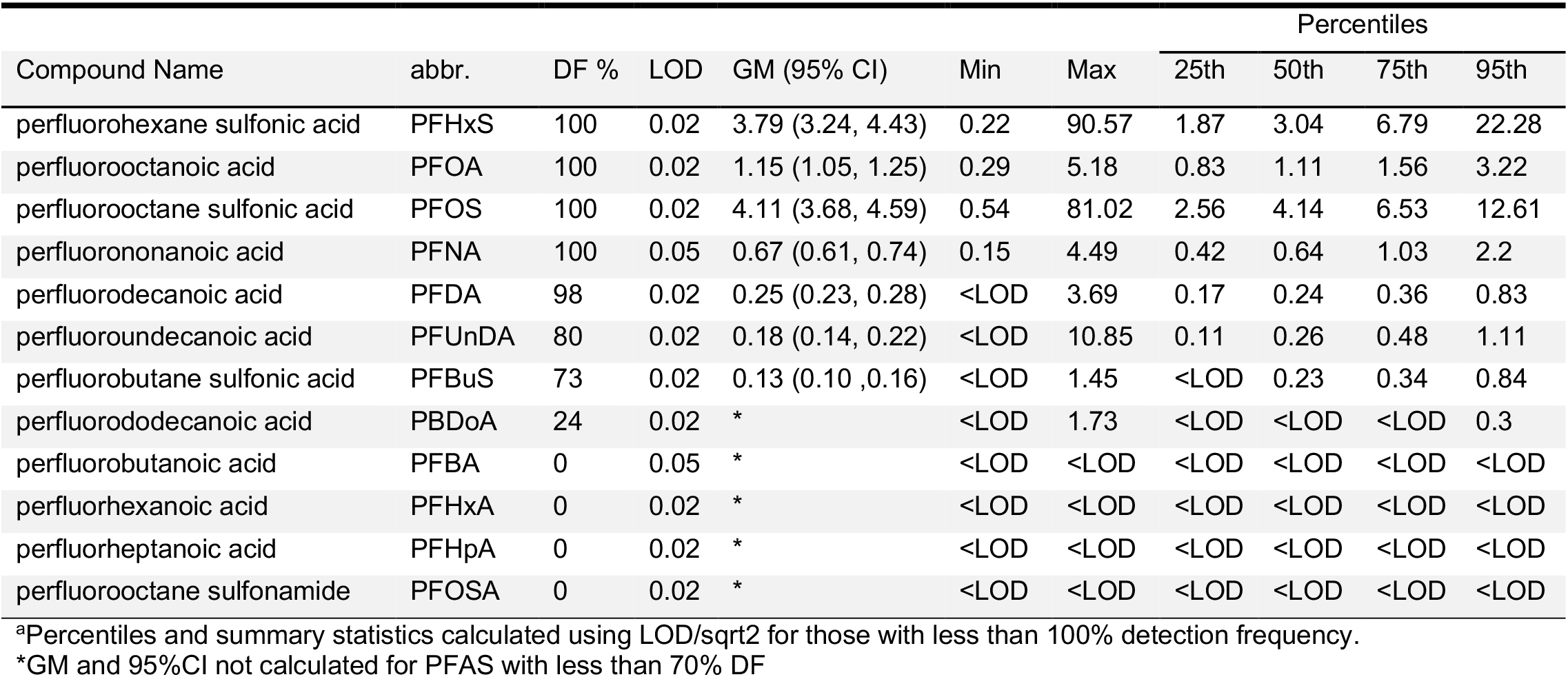
Geometric mean (GM) and 95% confidence interval (CI), detection frequency (DF), level of detection (LOD) (ng/mL) and percentiles^a^ for each PFAS analyzed in the WFBC cohort.

| Compound Name | abbr. | DF % | LOD | GM (95% CI) | Min | Max | Percentiles |  |  |  |
| --- | --- | --- | --- | --- | --- | --- | --- | --- | --- | --- |
|  |  |  |  |  |  |  | 25th | 50th | 75th | 95th |
| perfluorohexane sulfonic acid | PFHxS | 100 | 0.02 | 3.79 (3.24, 4.43) | 0.22 | 90.57 | 1.87 | 3.04 | 6.79 | 22.28 |
| perfluorooctanoic acid | PFOA | 100 | 0.02 | 1.15 (1.05, 1.25) | 0.29 | 5.18 | 0.83 | 1.11 | 1.56 | 3.22 |
| perfluorooctane sulfonic acid | PFOS | 100 | 0.02 | 4.11 (3.68, 4.59) | 0.54 | 81.02 | 2.56 | 4.14 | 6.53 | 12.61 |
| perfluorononanoic acid | PFNA | 100 | 0.05 | 0.67 (0.61, 0.74) | 0.15 | 4.49 | 0.42 | 0.64 | 1.03 | 2.2 |
| perfluorodecanoic acid | PFDA | 98 | 0.02 | 0.25 (0.23, 0.28) | <LOD | 3.69 | 0.17 | 0.24 | 0.36 | 0.83 |
| perfluoroundecanoic acid | PFUnDA | 80 | 0.02 | 0.18 (0.14, 0.22) | <LOD | 10.85 | 0.11 | 0.26 | 0.48 | 1.11 |
| perfluorobutane sulfonic acid | PFBuS | 73 | 0.02 | 0.13 (0.10, 0.16) | <LOD | 1.45 | <LOD | 0.23 | 0.34 | 0.84 |
| perfluorododecanoic acid | PBDoA | 24 | 0.02 | * | <LOD | 1.73 | <LOD | <LOD | <LOD | 0.3 |
| perfluorobutanoic acid | PFBA | 0 | 0.05 | * | <LOD | <LOD | <LOD | <LOD | <LOD | <LOD |
| perfluorhexanoic acid | PFHxA | 0 | 0.02 | * | <LOD | <LOD | <LOD | <LOD | <LOD | <LOD |
| perfluorheptanoic acid | PFHpA | 0 | 0.02 | * | <LOD | <LOD | <LOD | <LOD | <LOD | <LOD |
| perfluorooctane sulfonamide | PFOSA | 0 | 0.02 | * | <LOD | <LOD | <LOD | <LOD | <LOD | <LOD |

<sup>a</sup>Percentiles and summary statistics calculated using LOD/sqrt2 for those with less than 100% detection frequency.
\*GM and 95%CI not calculated for PFAS with less than 70% DF

**Table 4** shows the results from multiple linear regression models assessing the relationship between log-transformed PFAS concentrations and occupation, comparing firefighters to office workers (referent) controlling for potential confounders (**Tables S1 and S2**). Multiple regression models found higher serum levels of PFHxS, PFUnDA and PFNA among firefighters compared to office workers in both unadjusted in adjusted models. PFHxS levels were 2.39 (95%CI = 1.64,3.48) times higher, PFUnDA levels were 2.32 (95%CI = 1.17, 4.62) times higher and PFNA levels were 1.26 (95% CI = 0.99, 1.59) times higher in firefighters compared to office workers after adjusting for age, race and ethnicity, educational attainment, as well as the frequency of consumption of fish/shellfish, red meat, poultry, fast-food, food heated in packaging.

**Table 4.** Unadjusted and adjusted^a,b^ proportional change in geometric mean (95% confidence interval) of PFAS concentrations by firefighter status

| model | βCoef | expβ (95% CI) | p-value |
| --- | --- | --- | --- |
| **Perfluorononanoic Acid PFNA** |  |  |  |
| unadjusted | 0.23 | **1.25 (1.03, 1.53)<sup>†</sup>** | 0.0245 |
| model 1<sup>a</sup> | 0.20 | **1.23 (0.98, 1.54)\*** | 0.0794 |
| model 2<sup>b</sup> | 0.23 | **1.26 (0.99, 1.59)\*** | 0.0575 |
| **Perfluorooctanoic Acid PFOA** |  |  |  |
| unadjusted | -0.04 | 0.96 (0.81, 1.13) | 0.6070 |
| model 1<sup>a</sup> | 0.03 | 1.03 (0.85, 1.25) | 0.7364 |
| model 2<sup>b</sup> | 0.11 | 1.12 (0.91, 1.37) | 0.2837 |
| **Perfluorooctanoic Sulfonate PFOS** |  |  |  |
| unadjusted | 0.09 | 1.10 (0.88, 1.36) | 0.4097 |
| model 1<sup>a</sup> | 0.18 | 1.19 (0.93, 1.53) | 0.1621 |
| model 2<sup>b</sup> | 0.12 | 1.13 (0.87, 1.47) | 0.3623 |
| **Perfluorohexane Sulfonate PFHxS** |  |  |  |
| unadjusted | 0.43 | **1.54 (1.14, 2.07) <sup>†</sup>** | 0.0051 |
| model 1<sup>a</sup> | 0.67 | **1.96 (1.38, 2.78) <sup>†</sup>** | 0.0002 |
| model 2<sup>b</sup> | 0.87 | **2.39 (1.64, 3.48) <sup>‡</sup>** | 0.0000 |
| **Perfluorodecanoic Acid PFDA** |  |  |  |
| unadjusted | 0.13 | 1.14 (0.92, 1.43) | 0.2324 |
| model 1<sup>a</sup> | 0.13 | 1.14 (0.88, 1.49) | 0.3183 |
| model 2<sup>b</sup> | 0.16 | 1.18 (0.89, 1.55) | 0.2427 |
| **Perfluorobutane Sulfonate PFBuS** |  |  |  |
| unadjusted | 0.17 | 1.19 (0.65, 2.18) | 0.5731 |
| model 1<sup>a</sup> | 0.59 | 1.80 (0.87, 3.70) | 0.1129 |
| model 2<sup>b</sup> | 0.55 | 1.74 (0.81, 3.75) | 0.1580 |
| **Perfluoroundecanoic Acid PFUnDA** |  |  |  |
| unadjusted | 0.55 | **1.74 (1.00, 3.02) <sup>†</sup>** | 0.0496 |
| model 1<sup>a</sup> | 0.77 | **2.17 (1.10, 4.25) <sup>†</sup>** | 0.0245 |
| model 2<sup>b</sup> | 0.84 | **2.32 (1.17, 4.62) <sup>†</sup>** | 0.0162 |

<sup>a</sup> model 1 adjusted for: age, race/ethnicity, and education.
<sup>b</sup> model 2 adjusted for covariates in model 1 and consumption of: fish/shellfish, red meat, poultry, fast-food or take-out food, frozen food heated in paper or cardboard package. Office worker is referent group. \*pvalue ≤ 0.1; <sup>†</sup>pvalue ≤ 0.05; <sup>‡</sup>pvalue ≤ 0.001

When limiting our analysis to firefighters, we found that assigned firefighter position was associated with higher levels of six of the seven PFAS (**Table 5**). Having the occupational position of firefighter or officer (versus driver) was associated with higher average serum levels of PFNA, PFOA, PFOS, PFDA and PFUnDA, while drivers had higher average levels of PFBuS compared to those in officer and firefighter positions. Firefighters assigned to the airport had higher levels of PFNA than those assigned to other stations in the fire department. Likewise, firefighters who reported using firefighting foam had elevated PFOA levels compared to those who reported never using foams.

**Table 5.**
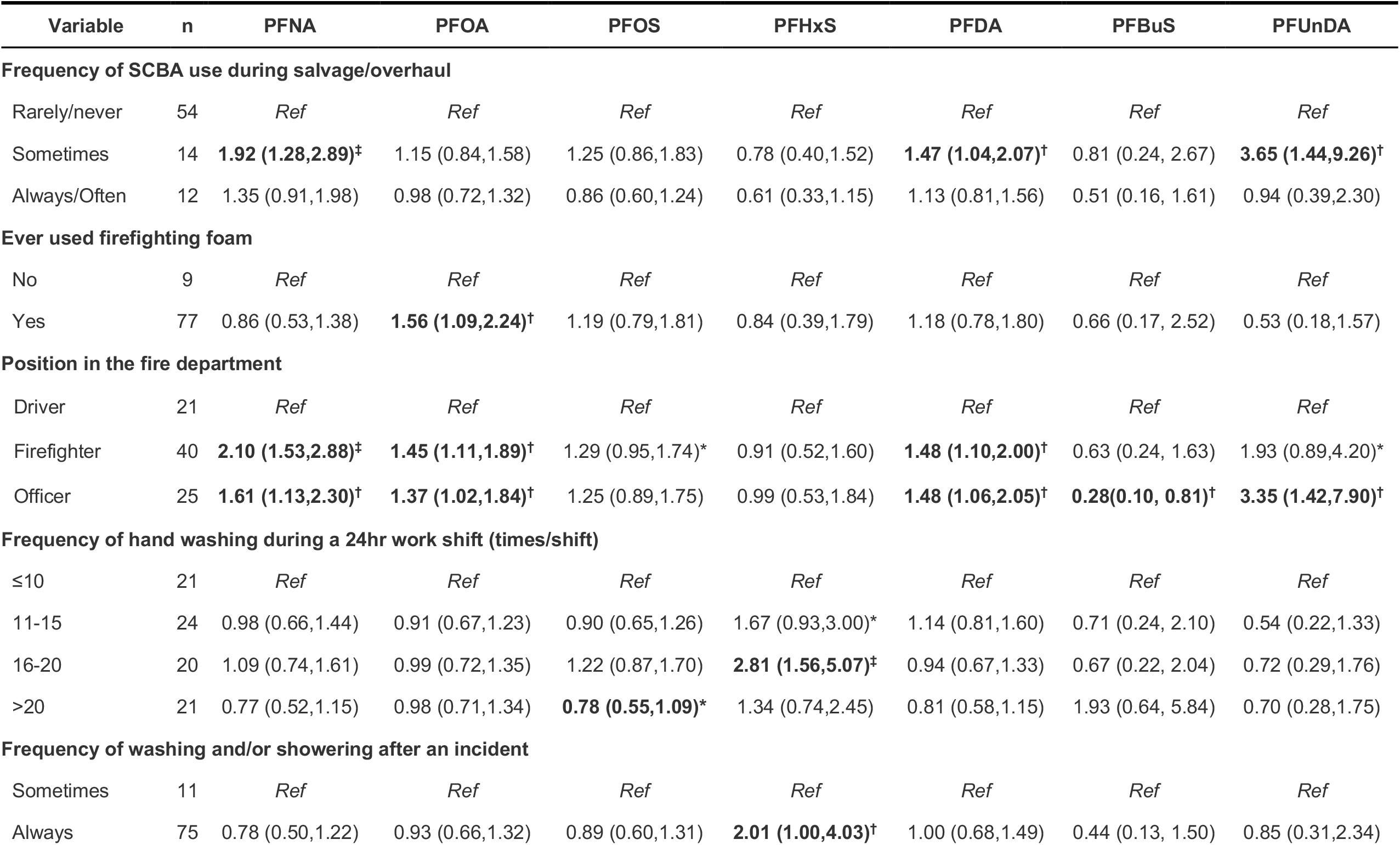
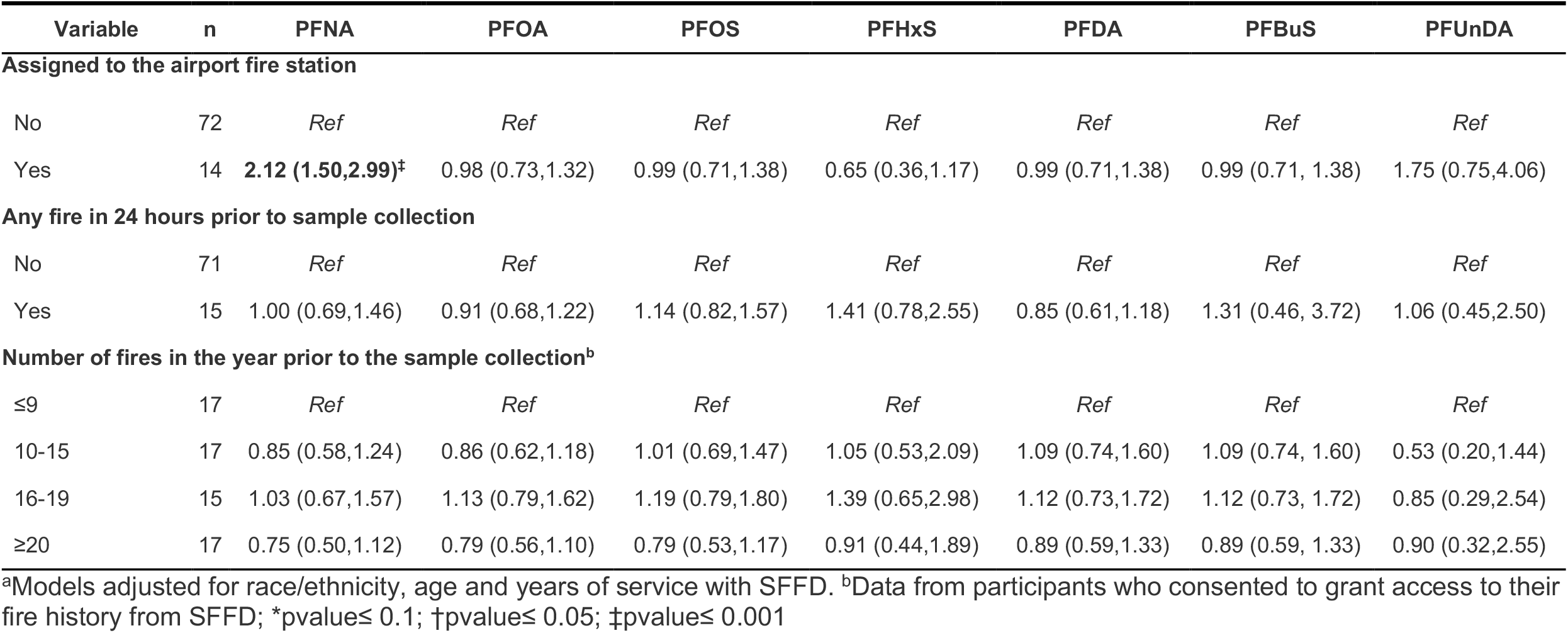
Adjusted proportional change in geometric mean (GM) and 95% confidence interval (CI) of covariates and serum PFAS concentration among firefighters (N=86) from Maximum Likelihood Regression (MLE) models^a^

Surprisingly, using SCBA, washing hands during a work shift and showering after a fire incident were associated with increased levels of some PFAS. Average PFNA and PFUnDA concentrations were higher among participants who said they sometimes used their SCBA during salvage/overhaul versus rarely or never. Similarly, average PFHxS levels were higher among participants who said they washed their hands more frequently or who responded that they always showered after a fire event compared to those who washed hands or showered after a fire event the least often. Those who washed their hands 16-20 times during a work shift had 2.81 (95%CI = 1.56, 5.07) times higher concentration of PFHxS compared to those who reported washing their hands 10 times or less per shift. Those who reported always showering after a fire event had 2.01 (95% CI = 1.0, 4.03) times higher PFHxS levels compared to those who reported that they sometimes showered.

**Figure 1** compares PFAS levels measured in our WFBC participants with other cohorts, including adult women from the 2013-2014 NHANES^54^ and the predominantly male 2010-2011 FOX study.^53^ WFBC firefighters and office workers had higher serum levels of PFHxS and a higher detection frequency for PFBuS than both NHANES and FOX; while PFBuS had low detection frequencies in both the FOX and NHANES studies (DF: 6.9% and 0.7%, respectively) the WFBC had a PFBuS detection frequency of 74% for firefighters and 70% for office workers. WFBC firefighters also had higher levels of PFNA, PFDA and PFUnDA levels compared to NHANES, whereas office workers had similar levels to NHANES for PFOS, PFOA and PFNA. Both WFBC firefighter and office worker groups had lower levels of PFOS, PFOA, PFNA, PFDA, and PFUnDA than those measured in the FOX study.

**Figure 1.**
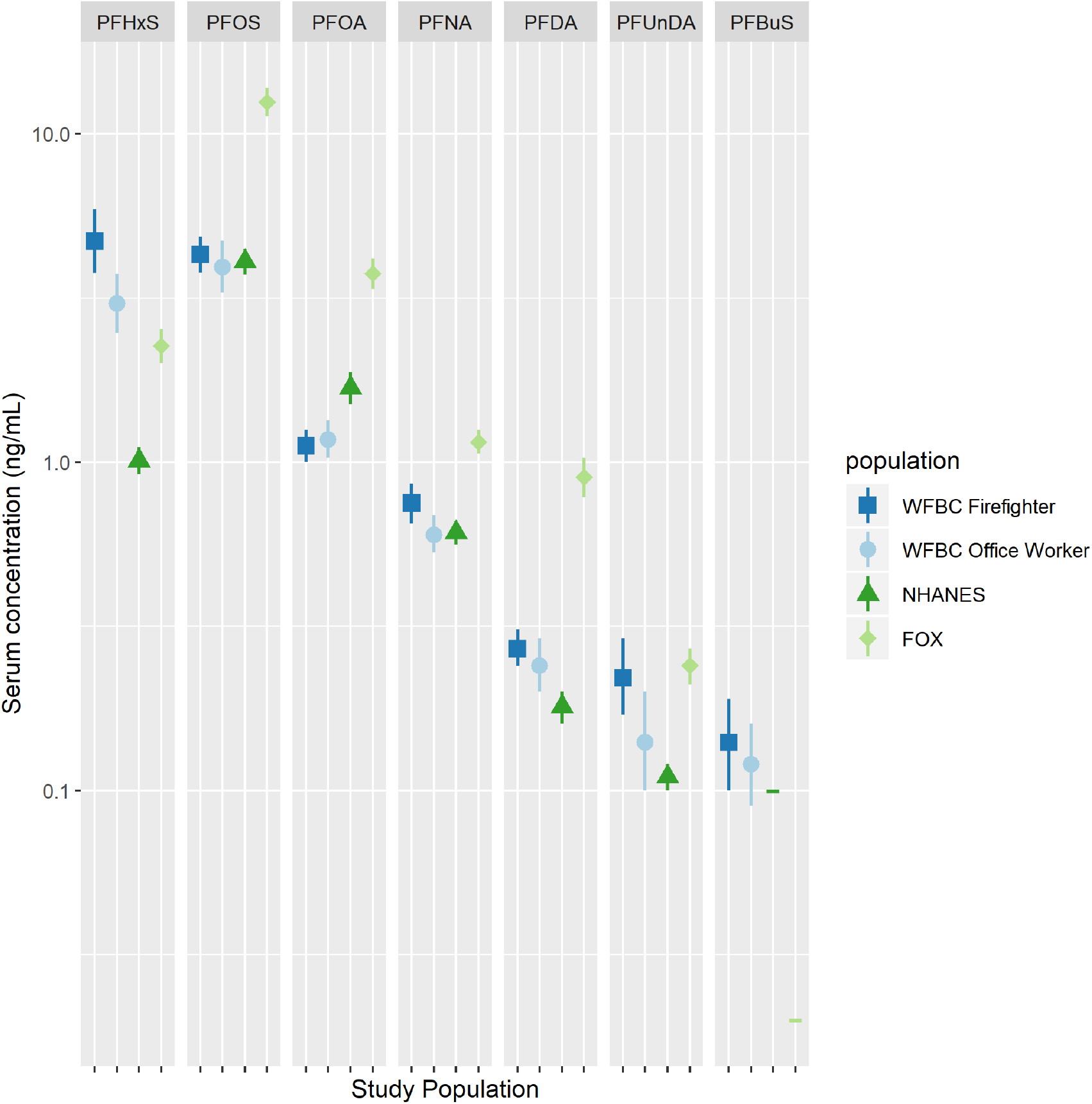
Comparison of geometric mean (GM) and 95% confidence intervals (CI) of PFAS levels measured in WFBC firefighters and office workers (2014-15) with adult women in NHANES (2013-14) and the FOX study (2010-11). Figure footnote: Figure shows GM and 95% CI for those PFAS with detection frequencies of at least 70% among WFBC participants. PFBuS had low DF for FOX (6.9%) and NHANES (0.7%); therefore, we plotted the LOD, represented by horizontal green lines, in lieu of GM and 95% CI. We replaced levels below the LOD with LOD/v2 and applied the LOD respective to each study population. Tests of statistical significance for differences between WFBC groups is shown in Table 4.

## Discussion

This biomonitoring study is the first to assess levels of PFAS compounds in an all-female cohort of firefighters and office workers. Of the twelve PFAS we measured, detection frequencies ranged from 0 to 100% and all participants had at least four PFAS congeners (PFHxS, PFOA, PFOS and PFNA) detected in their serum samples. Widespread use of PFAS in consumer products, contamination of food and water sources, and their environmental persistence may contribute to the high background levels of these compounds in both firefighters and office workers. Indeed PFAS are ubiquitous in the environment and have been found in dust, food and humans worldwide.^34,57^ In addition, the widespread use of PFAS containing products on clothing, furniture fabrics, carpets and paper food packaging also contributes to levels found in people.^48,58^ Drinking water contamination is considered an important source of PFAS exposure in many communities,^48,59,60^ and people with contaminated drinking water have elevated PFAS in their blood.^61^ However, local San Francisco Bay Area water systems are not likely to be a significant source of PFAS exposure for WFBC study participants. Municipal water systems in the locations where most study participants live were tested for PFAS under the Environmental Protection Agency’s Unregulated Contaminant Monitoring Rule in 2016 and tests did not detect measurable PFAS contamination in San Francisco or surrounding communities’ municipal water systems.^62^

Multivariate models showed that PFHxS, PFUnDA, and PFNA exposure was higher in firefighters compared to office workers after controlling for age, race and ethnicity. When we additionally controlled for the frequency of eating certain foods the association remained significantly higher in firefighters compared to office workers and the strength of association became stronger after adjustment for potential confounders. When we limited the analysis to firefighters, we found that several occupational activities were associated with higher PFAS levels. Firefighters’ assigned position was the most strongly associated with higher PFAS levels, with those assigned as firefighters or officers having higher levels of PFNA, PFOA, PFDA and PFUnDA compared to drivers. Compared to drivers, PFOS was higher among those assigned to the firefighter position however, PFBuS was higher among drivers compared to officers. Differential PFAS exposures by position may be explained by different roles at fire events. Engine drivers typically remain with the apparatus because they supply water for initial interior fire suppression work, as well as set the ladders for ventilation procedures. In addition, drivers in SFFD do not typically perform overhaul procedures after a fire is extinguished, and thus may avoid a critical source of exposure.^63^

Firefighting foams may be another important source of PFAS exposure. Aqueous Film Forming Foams (AFFF) are known to contain PFAS surfactants, including PFOS, PFOA and PFHxS.^19,64^ Our data suggest that firefighters who reported using firefighting foam had higher levels of PFOA than those who reported never using firefighting foams. Among airport firefighters, we found that PFNA levels were two times higher compared to firefighters assigned to other stations in San Francisco. Although PFNA is not considered a main ingredient in AFFF, a study in Finland found that firefighters using AFFF in training activities had increased levels of PFNA after the training activities.^19^

Unexpectedly, procedures that generally are intended to reduce contaminant exposures were associated with increased exposures for some PFAS. Washing hands more frequently during the work shift and always showering after a fire event were associated with increased PFHxS levels while sometimes using SCBA during salvage overhaul was associated with increased PFNA, PFDA and PFUnDA. One possible explanation is that firefighters may be over-reporting their hand washing, showering frequency, and SCBA use because they know that these activities are expected occupational hygiene practices. Duration of SCBA use during fire events may also be an important factor to examine in future studies, as it is possible that although firefighters may wear SCBA frequently, they may remove it after extinguishing a fire and during salvage and overhaul operations because gear is heavy, conditions are hot, and the location appears to be free of smoke.^14,63^ Self-reporting bias in terms of occupational safety and health procedures, which has been shown in other studies settings,^65,66^ may have also affected our results. In addition, there may be unknown and unmeasured occupational sources of PFAS exposure in firefighting that could help explain observed associations.

We compared the levels of PFAS congeners in WFBC participants with levels from two other exposure studies: A cohort of mostly male firefighters in Southern California (FOX) and a nationally representative sample of adult women (NHANES). In general, WFBC firefighters and office workers had lower PFAS levels than those measured in the FOX study, except for PFHxS which was significantly higher in both WFBC firefighters and office workers. In particular, the lower levels of PFOA and PFOS in the WFBC participants may reflect the temporal trends associated with the phase-out of these compounds in consumer products and firefighting equipment. The FOX study samples were collected between 2010 and 2011, three years before the start of the WFBC study and during the phase out period of PFOA and PFOS.^67^ Studies have shown that PFAS levels have changed over time with many PFAS that have been phased out decreasing and their replacements increasing.^36,37,68,69^ Interestingly, we did see overall higher levels of PFHxS in both groups of WFBC participants collected in 2013-2014 relative to the 2010-2011 FOX cohort. The increased levels in WFBC participants may be due to PFHxS’ long half-life or other continued sources of PFHxS exposures in California residents.^68^ The elevated levels among WFBC firefighters in relation to FOX participants may be due, in part, to its use as a replacement for PFOA and PFOS in firefighting foams.^64^ Additionally, the lower levels of most PFAS among WFBC participants compared to the mostly male cohort of FOX participants may be further explained by excretion pathway differences between men and women, where women may have lower PFAS levels due to excretion during menstruation.^70^

WFBC and NHANES data were collected at around the same time (2013-14 for NHANES and 2014-15 for the WFBC) and levels between WFBC and NHANES were more similar to each other than with the FOX study. However, PFHxS, PFDA, and PFBuS levels were higher in WFBC firefighters and office workers compared to NHANES adult women. While we limited the NHANES cohort to adult women, we did not consider occupation or other factors, including location or other potential exposure sources.

Health effects observed in people exposed to PFOA include high cholesterol, ulcerative colitis, thyroid disease, testicular and kidney cancers, and pregnancy-induced hypertension.^71^ PFOS exposure has also been associated with immunotoxicity, as indicated by a decreased response to vaccine in children;^72^ and other studies of people exposed to PFAS show effects on liver and decreased birth weight.^73^ In animal studies, PFAS have been shown to have a variety of similar toxicological effects including liver toxicity, suppressed immune function, altered mammary gland development, obesity, and cancer.^74,75^ The concordance between endpoints identified in animal studies and human studies, most notably effects on liver, kidney, fetal growth and development, and suppression of the immune system, add confidence to the findings.^76,77^ Based on these studies in humans and animals, some government agencies have established allowable levels of PFOA and PFOS in drinking water.^76,78^ For PFOA, these benchmarks are designed to prevent people from having blood serum levels above 14 ng/ml.^79^ The maximum value in this study was 5 ng/mL. However, PFOA has been shown to alter mammary gland development in mice following in utero exposure at even lower levels, and estimates of target serum levels to protect against those effects are less than 1 ng/ml^79^. The health effects of the other PFAS found in elevated levels in firefighters have not been well studied and target serum concentrations that are expected to protect against adverse effects are not available for PFNA, PFBuS, PFHxS, PFUnDA, so it is difficult to compare measured levels in firefighters with benchmarks intended to protect against health effects. Allowable daily intake amounts intended to protect against adverse health effects are in the same range as PFOA for PFNA, while for PFHxS they are about 10 times higher and for PFBuS they are 100 times higher.^80^ However the higher allowable daily intakes for PFBuS and PFHxS do not necessarily correspond to higher allowable serum levels for these chemicals. Instead they reflect differences in the relationship between intake and serum concentration for each of the chemicals.

We used a community-based participatory research design that entailed the active involvement of firefighters to recruit participants (both firefighters and office workers) into the study. By including office workers from the City and County of San Francisco we were able to compare firefighter exposures to a working population of women who were not involved in firefighting activities, but who live and work in the same geographical region allowing us to examine which PFAS chemicals are most strongly associated with firefighting. Understanding the extent to which PFAS exposures differ between firefighters and office workers can elucidate which compounds are likely to have occupational sources, highlight opportunities for prevention strategies, and assess the effectiveness of workplace exposure reduction efforts.

## Conclusion

Our study found ubiquitous exposures to multiple PFAS compounds among all participants of the Women Firefighter Biomonitoring Collaborative. Firefighters were exposed to higher levels of PFNA, PFHxS and PFUnDA than office workers indicating potential occupational sources for firefighters. While industrial hygiene practices, use and access to personal protective equipment, use of AFFF foam, and other occupational conditions may vary across fire departments and fire events, our results have relevance for understanding potential sources and pathways of chemical exposures for other fire departments and regions. Further study is warranted to better understand exposure patterns and health risks for firefighters associated with these compounds.

## Data Availability

Data and code will be made available post-publication.

## Acknowledgements

The authors thank all of the WFBC study participants for their contribution to the study. This work is supported by the California Breast Cancer Research Program #19BB-2900 (JT, RG, TL, RAR, HB, VB, RMF), the National Institute of Environmental Health Sciences R01ES027051 (RMF) and the San Francisco Firefighter Cancer Prevention Foundation (HB). We thank Anthony Stefani, Cassidy Clarity, Emily O’Rourke, Nancy Carmona, Karen Kerr, Julie Mau, Natasha Parks, Lisa Holdcroft, San Francisco Fire Chief Jeanine Nicholson, former San Francisco Fire Chief Joanne Hayes-White, Sharyle Patton, Connie Engel and Nancy Buermeyer for their contributions to the study.

RAR and VB, are employed at the Silent Spring Institute, a scientific research organization dedicated to studying environmental factors in women’s health. The Institute is a 501(c)3 public charity funded by federal grants and contracts, foundation grants, and private donations, including from breast cancer organizations. HB is former president and member of United Fire Service Women, a 501(c)3 public charity dedicated to supporting the welfare of women in the San Francisco Fire Department.

The authors declare they have no actual or potential competing financial interests.

